# Development of a pilot machine learning model to predict successful cure in critically ill patients with community-acquired pneumonia

**DOI:** 10.1101/2025.07.14.25331407

**Authors:** Mengou Zhu, Wan-Ting Liao, Alec Peltekian, Nikolay S. Markov, Mengjia Kang, Luke V. Rasmussen, Thomas Stoeger, Theresa L. Walunas, Alexander V. Misharin, Benjamin D. Singer, G. R. Scott Budinger, Richard G. Wunderink, Ankit Agrawal, Catherine A. Gao

## Abstract

Severe community-acquired pneumonia (CAP) remains a major cause of critical illness, yet there are no validated early clinical criteria to predict short-term treatment outcomes in these patients. Short-term pneumonia treatment outcomes are less affected by confounding factors introduced by a prolonged hospital course, and early prediction of short-term treatment outcomes can help physicians identify those who are likely to fail the current treatment and implement adjustments to existing diagnostic and therapeutic plans. Traditional clinical stability criteria such as Halm’s criteria are not calibrated for early outcome prediction in critically ill severe pneumonia patients. We applied the XGBoost algorithm to predict pneumonia cure by day 7-8 post-intubation with clinical features from days 1-3 in mechanically ventilated patients with severe CAP from the Successful Clinical Response in Pneumonia Therapy (SCRIPT) study, a prospective cohort study at a tertiary academic center. Pneumonia episodes were adjudicated for day 7-8 cure status by a panel of critical care physicians using a structured review process. Clinical features that inform Halm’s criteria, including vital signs, oxygenation parameters, mental status, and vasopressor use, were extracted from the electronic health record. We also examined model performance by including additional features, such as laboratory data, ventilator settings, and medications. Basic demographic characteristics including age and BMI were also incorporated. Among 85 patients, 42 (49.4%) were cured by day 7–8. The best-performing model, which used Halm’s clinical features and ventilator features from days 1–3, achieved a cross-validated AUROC of 0.757. Inclusion of lab and medication data did not significantly improve performance. Key predictors included GCS, norepinephrine requirement, and BMI. We prove the feasibility of using ML models to predict short-term treatment outcomes of severe CAP among critically ill patients with basic clinical features. Future studies should focus on external validation and clinical integration to inform prognosis and early reevaluation of treatment strategy in patients with predicted poor outcomes.

## Background

Pneumonia is a leading cause of hospitalizations and mortality in the United States.^1^ Early, accurate outcome prediction is crucial for identifying patients who are likely to fail pneumonia treatment, which allows for timely adjustments to patient management strategies, especially in critical care settings. Currently, there are no established clinical criteria to predict pneumonia cure using early clinical features in critically ill patients with severe community-acquired pneumonia (CAP). Clinical stability criteria, such as Halm’s Criteria (based on vital signs, mental status, and oral intake), measure and predict clinical stability and are suitable for identifying candidates for antibiotic de-escalation and hospital discharge.^2^ These are not suitable, however, for prediction of CAP cure in critically ill patients with severe pneumonia.^3^ Previous attempts to use machine learning (ML) methods to predict pneumonia treatment outcome using clinical features focus on long-term outcomes such as 30-day mortality, which are likely to be confounded by factors other than pneumonia cure.^45^ We hypothesized that machine learning based on features from early days following intubation predict short-term treatment outcomes in severe CAP.

## Methods

We analyzed CAP episodes from patients enrolled in the Successful Clinical Response in Pneumonia Therapy (SCRIPT) study, a prospective observational cohort study of patients receiving mechanical ventilation who underwent bronchoalveolar lavage (BAL) for evaluation of suspected pneumonia.^6–8^ Patient’s families or representatives consented to participate in the study, which was approved by the Northwestern University IRB STU00204868. A group of critical care attending physicians reviewed the charts and adjudicated whether the pneumonia episode was successfully cured at day 7-8 after BAL using a previously-described, multiple independent reviewer process.^8^ Basic demographic features including age and BMI, as well as clinical features from days 1-3 of each CAP episode were used to build several XGboost^9^ ML models to predict whether the pneumonia episode would successfully be treated (cured) by day 7-8 (Figure 1A). The clinical features of interest were derived from the original Halm’s Criteria, including the daily average and/or worst values of temperature, heart rate, systolic blood pressure, respiratory rate, oxygen saturation (SpO2), and partial pressure of oxygen in arterial blood (PaO2). To account for life support interventions that may interfere with these variables, we also included fraction of inspired oxygen (FiO2), PaO2/FiO2 ratio, and norepinephrine rate. RASS and GCS were included as measures of mental status. Oral intake data was not readily available in the variables we pulled, but enteral nutrition was administered via orogastric tube for most patients. We compared our model performance with inclusion or exclusion of medication use, including steroid dose on the day(s) of assessment, cumulative steroid dose, cumulative Narrow-Spectrum Antibiotic Treatment (NAT) score, and the use of remdesivir. Similarly, we also compared inclusion and exclusion of laboratory parameters and ventilator parameters. XGBoost can inherently handle missing data, so no imputation was performed. We performed an 80:20 train/test split, and 10-fold cross validation on the training set. All splits/folds were performed at the patient level. SHAP plots were used for model interpretability. Hyperparameter tuning was performed using Optuna^10^. We evaluated models using AUROC.

**Figure 1.**
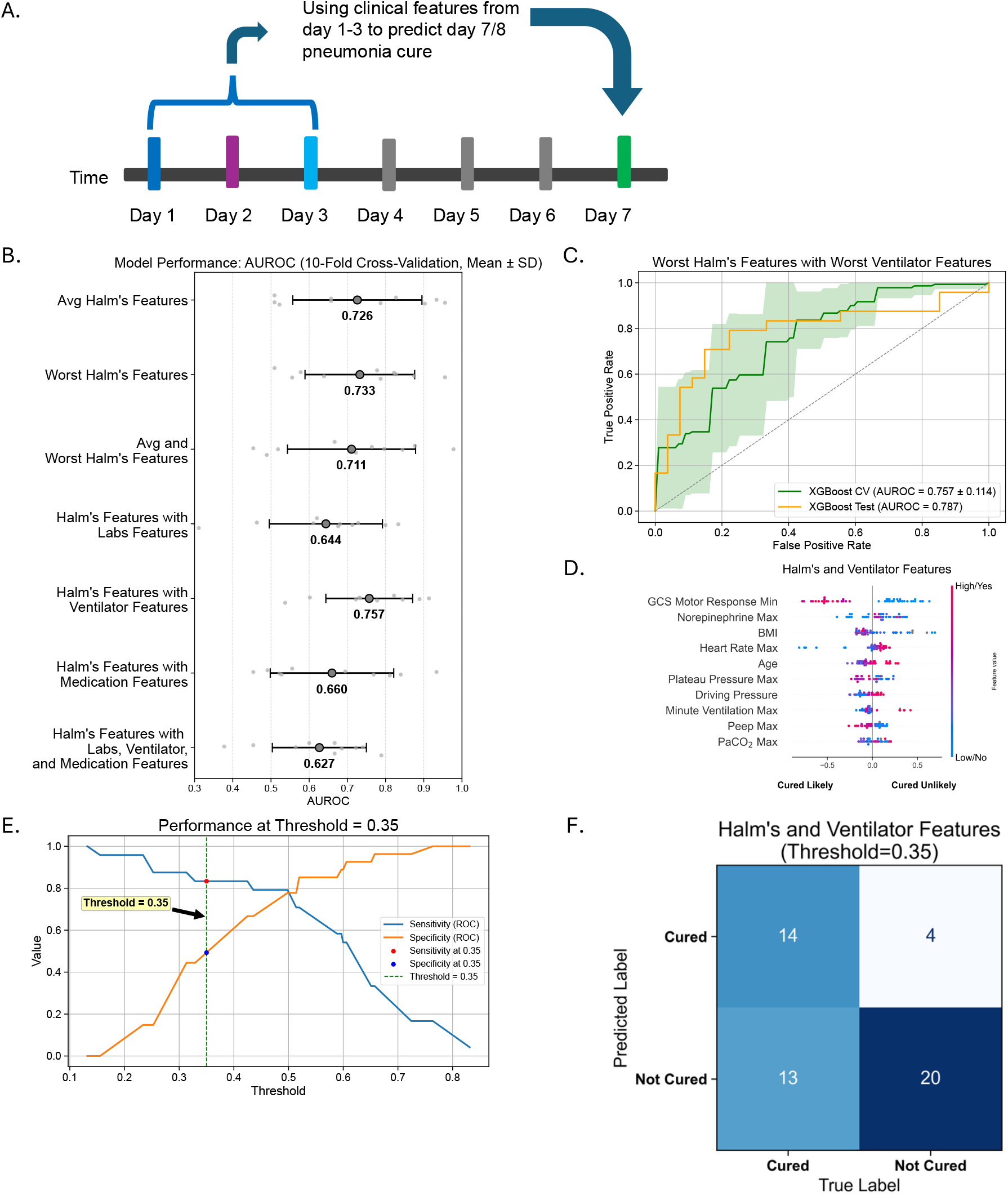
**A**. Schematic of using early clinical features to predict short-term pneumonia treatment outcome. **B**. 10-fold cross-validated AUROC of each XGBoost model tested, with the best-performing model being XGBoost with worst Halm’s features. **C**. AUROC of 10-fold cross-validation (green shading indicating average AUROC) and test set performance of XGBoost with worst Halm’s features and XGBoost with worst features and worst labs. **D**. SHAP plots indicating important features of XGBoost with worst Halm’s features and XGBoost with worst features and labs. **E**. Sensitivity and specificity at different thresholds, with chosen threshold at 0.35. **F**. Confusion matrix at threshold 0.35.

## Results

We examined ICU data from 85 patients with CAP that had clinical data from the first three episode days and an adjudicated day 7-8 cure status, out of which 42 (49.4%) were cured by day 7-8. 255 ICU days were included in the analysis. Patient demographics and outcomes are summarized in Table 1. None of the patients met Halm’s criteria in their first 3 days. Out of all the models assessed, the best-performing model was XGBoost that included the worst Halm’s clinical features and worst ventilator features from days 1-3, achieving a 10-fold cross validated AUROC of 0.757 (Figure 1B). This best-performing model achieved an AUROC of 0.787 on the test set (Figure 1C). Adding medications and laboratory parameters did not significantly improve model performance (Figure 1B). Important features factored into prediction included GCS, norepinephrine use, and BMI (Figure 1D). Model threshold was selected to prioritize sensitivity in identifying patients likely to fail treatment. At a threshold of 0.35, the model achieves a sensitivity of 0.83 and specificity of 0.52 in identifying patients who are likely to fail pneumonia treatment within the next few days (Figure 1E). The confusion matrix corresponding to this threshold is presented in Figure 1F.

**Table 1.** **A**. Demographics and outcomes for CAP cohort. Patient characteristics stratified by Day 7 episode resolution (Cured vs. Not Cured). Continuous variables are presented as median [first quartile, third quartile]; categorical variables are presented as number (percentage). BMI = Body Mass Index; SOFA = Sequential Organ Failure Assessment; NAT = Narrow Antibiotic Therapy; LTACH = Long-Term Acute Care Hospital; SNF = Skilled Nursing Facility; ECMO = Extracorporeal Membrane Oxygenation. **B**. Summary of the average and worst Halm’s clinical features from pneumonia episode day 1-3. Data presented in the format of median [Q1, Q3].

|  | Total | Cured | Not Cured |
| --- | --- | --- | --- |
| Number | 85 | 42 | 43 |
| Age (years), median [Q1,Q3] | 66.0 [55.0,75.0] | 65.0 [52.8,73.0] | 67.0 [55.0,75.0] |
| Female, n (%) | 42 (49.4) | 21 (50.0) | 21 (48.8) |
| Body Mass Index (kg/m2), median [Q1,Q3] | 27.7 [22.8,33.5] | 27.9 [23.8,33.9] | 25.8 [21.5,33.1] |
| Immunocompromised, n (%) | 10 (11.8) | 8 (19.0) | 2 (4.7) |
| SOFA Score on Episode Day 1, median [Q1,Q3] | 12.0 [10.0,15.0] | 12.0 [9.0,13.8] | 12.0 [11.0,15.0] |
| Transferred from External Facility, n (%) | 6 (7.1) | 3 (7.1) | 3 (7.0) |
| Race, n (%) |  |  |  |
| White | 47 (55.3) | 22 (52.4) | 25 (58.1) |
| Black or African American | 24 (28.2) | 12 (28.6) | 12 (27.9) |
| Asian | 2 (2.4) | 1 (2.4) | 1 (2.3) |
| Unknown or Not Reported | 12 (14.1) | 7 (16.7) | 5 (11.6) |
| Ethnicity, n (%) |  |  |  |
| Hispanic or Latino | 12 (14.1) | 8 (19.0) | 4 (9.3) |
| Not Hispanic or Latino | 70 (82.4) | 33 (78.6) | 37 (86.0) |
| Unknown or Not Reported | 3 (3.5) | 1 (2.4) | 2 (4.7) |
| Discharge Disposition, n (%) |  |  |  |
| Home | 22 (25.9) | 18 (42.9) | 4 (9.3) |
| Rehab | 18 (21.2) | 13 (31.0) | 5 (11.6) |
| LTACH | 7 (8.2) | 2 (4.8) | 5 (11.6) |
| SNF | 11 (12.9) | 8 (19.0) | 3 (7.0) |
| Hospice | 3 (3.5) |  | 3 (7.0) |
| Died | 24 (28.2) | 1 (2.4) | 23 (53.5) |
| Received ECMO during Episode, n (%) | 17 (20.0) | 6 (14.3) | 11 (25.6) |
| Underwent Tracheostomy, n (%) | 9 (10.6) | 2 (4.8) | 7 (16.3) |
| Total Hospitalization Days, median [Q1,Q3] | 18.0 [10.0,27.0] | 18.0 [12.0,24.8] | 16.0 [7.5,30.0] |
| Total ICU Days, median [Q1,Q3] | 9.0 [6.0,15.0] | 8.0 [6.2,11.8] | 11.0 [6.0,20.0] |
| Episode Duration, median [Q1,Q3] | 7.0 [4.0,7.0] | 5.0 [4.0,7.0] | 7.0 [5.5,12.0] |

B.
|  | Day 1 | Day 2 | Day 3 |
| --- | --- | --- | --- |
| **Temperature, average** | 98.3 [97.7,99.5] | 98.3 [97.9,99.3] | 98.4 [98.2,99.2] |
| **Temperature, worst** | 99.7 [98.8,101.3] | 99.4 [98.8,100.6] | 99.5 [98.9,100.8] |
| **Heart rate, average** | 89.2 [79.5,102.0] | 82.9 [70.4,93.3] | 80.7 [71.1,93.2] |
| **Heart rate, worst** | 113.0 [94.0,130.0] | 100.0 [87.0,116.0] | 100.0 [89.0,121.0] |
| **Systolic blood pressure, average** | 112.2 [104.5,122.0] | 113.8 [104.9,124.4] | 116.3 [109.0,128.6] |
| **Systolic blood pressure, worst** | 82.0 [79.0,89.0] | 87.0 [79.0,98.0] | 91.0 [82.0,101.0] |
| **Diastolic blood pressure, average** | 59.8 [56.3,66.5] | 57.5 [53.9,62.7] | 60.5 [54.5,65.1] |
| **Diastolic blood pressure, worst** | 46.0 [41.0,50.0] | 46.0 [42.0,50.0] | 47.0 [43.0,50.0] |
| **Respiratory rate, average** | 22.8 [19.5,26.0] | 21.6 [18.0,25.1] | 19.6 [17.2,24.3] |
| **Respiratory rate, worst** | 29.0 [23.0,35.0] | 26.5 [22.0,32.0] | 26.0 [22.0,32.0] |
| **Oxygen saturation, average** | 95.9 [94.3,97.0] | 96.6 [94.8,98.0] | 96.5 [94.9,98.0] |
| **Oxygen saturation, worst** | 92.0 [87.0,94.0] | 93.0 [90.0,96.0] | 93.0 [90.0,96.0] |
| **Norepinephrine, average** | 0.1 [0.0,0.2] | 0.0 [0.0,0.1] | 0.0 [0.0,0.1] |
| **Norepinephrine, worst** | 0.1 [0.0,0.3] | 0.1 [0.0,0.1] | 0.0 [0.0,0.1] |
| **RASS, average** | -2.9 [-4.0,-1.7] | -2.9 [-4.0,-1.3] | -2.0 [-3.8,-0.9] |
| **RASS, worst** | -4.0 [-5.0,-4.0] | -4.0 [-5.0,-3.0] | -3.0 [-5.0,-2.0] |
| **GCS (eye opening), average** | 2.0 [1.0,2.5] | 2.0 [1.0,3.0] | 2.8 [1.2,3.7] |
| **GCS (eye opening), worst** | 1.0 [1.0,2.0] | 1.0 [1.0,2.0] | 2.0 [1.0,3.0] |
| **GCS (motor response), average** | 3.2 [1.6,4.8] | 3.3 [1.0,5.0] | 4.0 [1.5,5.6] |
| **GCS (motor response), worst** | 1.0 [1.0,4.0] | 1.0 [1.0,4.0] | 3.0 [1.0,5.0] |
| **GCS (verbal response), average** | 1.0 [1.0,1.0] | 1.0 [1.0,1.0] | 1.0 [1.0,1.0] |
| **GCS (verbal response), worst** | 1.0 [1.0,1.0] | 1.0 [1.0,1.0] | 1.0 [1.0,1.0] |
| **FiO2, average** | 50.0 [41.1,64.7] | 40.0 [32.7,50.0] | 36.8 [30.0,45.7] |
| **FiO2, worst** | 80.0 [50.0,100.0] | 40.0 [40.0,60.0] | 40.0 [30.0,60.0] |
| **PaO2, average** | 104.3 [87.1,121.4] | 96.2 [80.3,118.9] | 92.8 [77.4,111.2] |
| **PaO2, worst** | 72.0 [61.0,84.5] | 77.0 [63.8,92.0] | 76.0 [65.0,91.5] |
| **P/F ratio, average** | 196.8 [160.5,276.3] | 249.6 [192.0,327.5] | 260.0 [189.1,322.1] |
| **P/F ratio, worst** | 151.7 [100.8,204.2] | 206.2 [145.6,283.8] | 204.0 [141.7,289.1] |

## Discussion

ML models provide promising results in predicting severe CAP short-term treatment outcomes. To our knowledge, this is the first attempt at making short-term ML treatment outcome prediction based on early clinical features among critically ill patients with severe CAP. Short-term pneumonia treatment outcomes are less affected by confounding factors introduced by a prolonged hospital course, and early prediction of short-term treatment outcomes can help physicians identify those who are likely to fail the current treatment and implement adjustments to existing diagnostic and therapeutic plans. Patients identified by the model to be at high risk of not being cured by day 7-8 may benefit from additional investigation with imaging, repeat BAL, or change in antibiotic regimen. Our study proves that such prediction is feasible. Careful adjudication of pneumonia cure by a panel of physician experts is a strength of our study. Limitations include sample size and recruitment from a single tertiary care academic medical center with standard of care BAL to guide antibiotic management. We are unaware of the existence of another dataset that has this level of annotation, and therefore lack an external validation dataset. Future studies can prospectively evaluate the performance of our model by adjudicating pneumonia outcomes using published criteria.

## Data availability

An earlier version that includes the majority of this cohort is available to credentialed users who sign a DUA at https://physionet.org/content/script-carpediem-dataset/1.1.0/ and an update with new patients enrolled since publication of the initial dataset is under preparation.

## Code availability

Code for processing and reproducing our analysis is available at https://github.com/NUSCRIPT/mz_pna_cure_2025.

Human Ethics and Consent to Participate: This study was approved by the Northwestern University IRB STU00204868. Patients’ families or representatives consented to partake in the study.

## Author contributions

Conceptualization: CAG

Methodology: MZ, WL, RGW, AA, CAG

Data acquisition: WL, MK, NM

Programming/analysis/visualization: MZ, WL, AP, NM, CAG

Drafting: MZ, WL, CAG

Editing and approval: all authors

